# Development of a women empowerment framework to promote voluntary medical male circumcision uptake for HIV prevention in Zimbabwe

**DOI:** 10.1101/2021.08.25.21262388

**Authors:** Grace Danda, Thandisizwe R Mavundla, Christina Mudokwenyu- Rawdon

## Abstract

**Introduction:** This study aimed to develop a women empowerment framework to promote Voluntary Medical Male Circumcision (VMMC) uptake for human immune-deficiency virus (HIV) prevention, guided by the following objectives: To explore relevant literature on the role of women in promoting male circumcision uptake; To develop a women empowerment framework to promote male circumcision uptake; To describe the women empowerment framework to promote male circumcision uptake.

**Methods:** The study used a theory-generation design to explore, describe and develop a women empowerment framework from a broad literature review.

**Results:** A three-phase approach suitable for the framework development emerged from the literature review. Phase one involved exploring relevant literature on the role of women in male circumcision uptake, using the Population/problem, Intervention, Comparison and Outcome (PICO) method to identify and define the key concepts. Phase 2 adopted and adapted the model by Maibvise and Mavundla in identifying the following concepts: change agents as the health care providers, recipients as the women to influence men eligible for male circumcision, promoting male circumcision uptake and success of the male circumcision program. Phase 3 provided a detailed description of the framework including three key steps of empowerment of women, influencing positive perceptions of men and utilisation of male circumcision services.

**Conclusions:** The identified concepts resulted in development of a women empowerment framework, which can be used as an education and advocacy tool in building the capacity of women in supporting the male circumcision programme to promote uptake. Further research is required to expand the utilisation of the framework in male circumcision healthcare services.

## INTRODUCTION

Since the introduction of VMMC in 2009, Zimbabwe has been struggling to meet its target of circumcising 80% of males, recommended by WHO to be achieved by year 2016 to facilitate HIV infection reduction by 3.4million in the 14 priority countries [1]. This is despite advancing various demand creation approaches put in place, such as media adverts, social mobilization in schools and communities and use of promotional materials. Barriers to men seeking circumcision include fear of pain, myths and misconceptions about infertility following circumcision. Some of the men perceived themselves as not being susceptible to HIV hence seeing no need to undergo the procedure while others who were not aware of their HIV status had fear of being tested for HIV as a barrier to VMMC uptake [2, 3]. Deterrents to VMMC such as lack of partner, parent or social support emerged in support of these factors [4].

The Ministry of Health (MOHCC) in Zimbabwe, in partnership with Non-Governmental organisations (NGOs) and private health sector, implement the VMMC programme using various approaches in demand creation. The MOHCC allocate each organization an operational setting for increased countrywide coverage and accountability, being mindful of the 80% target. Despite such a broad-based approach, Zimbabwe has struggled to meet the expected target, which was reset in 2016 to 90% of 15-29year olds in the priority countries by 2021 [5]. The 2018 country VMMC coverage among the target group of 15-29years was at 33% and the overall coverage of all circumcisions was at 68%, still falling short of the 80% overall target [6]. A multicountry study involving Tanzania, South Africa and Zimbabwe found that young females influenced partners’ decision to seek VMMC but influence of social network on VMMC uptake has not been fully explored [7,8]. In Zimbabwe and Tanzania, most of the women participating in the study felt that the indirect influence was through refusing to initiate an affair with a men or quickly discontinuing them if they refused circumcision. Some of the women however felt that men needed to be persuaded by being told about risks of infections like HIV and the cancers associated with not circumcising e.g. cervical cancer. Other participants thought that a woman should show her care for her man by convincing him to go for circumcision, regardless of the benefits she gets [7]. Older men have given reasons for not circumcising as partner refusal hence the need to increase awareness of VMMC among women [9]. There is no evidence in all these studies that a women empowerment framework was used to prepare women for their supportive role in VMMC, yet the framework is one part of the VMMC package that ensures women’s involvement in supportive care for men seeking circumcision services in Zimbabwe. Development of a framework to empower women using available literature was the main objective of this study. It can be used as an education and advocacy tool in building women’s capacities for effectiveness in their role as supportive female partners and mothers in the VMMC programme to promote uptake of male circumcision healthcare services.

## METHODS

### Study design

A theory generative design was used in this study in a bid to formulate a framework. Framework development involves identifying, relating and clarifying the relationships among concepts to show a true reflection representing the idea of focus [10]. The study was developed from the generated framework to enable the researcher to associate the results with nursing and public health knowledge building [11]. A women empowerment framework to promote the uptake of VMMC was developed from a broad literature search on the role of women in VMMC. The literature review and selection process was guided by PICO [12] to screen out studies that did not focus on the research question. Consultations with experts in VMMC and a local mentor, published research studies and the university supervisor were also utilised in the process.

In view of the fact that very few studies have been conducted on models and frameworks for improving VMMC uptake, the researcher employed a theory generative design to have a deeper understanding of women empowerment and VMMC uptake in order to develop a framework to promote VMMC uptake.

### Ethical considerations

The narrative and literature review did not involve human participants and therefore no ethical approval was needed for this study. However, the researchers were cleared by the Health Studies Research Ethics Committee (HSREC) of the University of South Africa (Unisa), the Mpilo Central Hospital ethics board and the Medical Research Council of Zimbabwe because this study was part of a PhD thesis. The scientific integrity of this research was enhanced by avoiding research misconduct through reporting the reviewed literature accurately, honestly and without distortions and all sources referred to were acknowledged in text and references to avoid plagiarism.

### Data collection

The narrative review of relevant literature was undertaken from January 2017 to January 2021. Studies relevant to male circumcision in HIV prevention dating from 2005 to 2020, published in English, were identified. This period was chosen to ensure broadness of period of studies, to facilitate adequate comparison and identify the most relevant and current concepts. Searches were made from Pub Med, Medline, Embase, CINHAL, MIDIRS, and Cochrane databases of systemic reviews (CDSR), Cochrane (Central) Register of Controlled Trials, www.whoint, Academia.edu and Google Scholar. The key words guiding literature review were women empowerment, framework, voluntary medical male circumcision, VMMC uptake and HIV prevention. Books, papers and studies reviewed were from UK, United States of America and the African region. Books and chapters were included as an initial source of providing an overview about HIV prevention strategies and male circumcision. Both narrative and systematic reviews were included as demonstrated in the following section.

## RESULTS

### PICO literature review

The review and selection process was guided by PICO [12] to screen out studies that did not focus on the research question. PICO is a literature search strategy used to select the appropriate relevant literature and to screen studies that do not match the criteria. This is an acronym standing for population of studies on role of women in VMMC; Intervention is the promotion of VMM uptake; Comparison is looking at barriers of VMMC causing reduced uptake and Outcome focuses on benefits of VMMC acting as facilitators to VMMC uptake.

**Fig 1:**
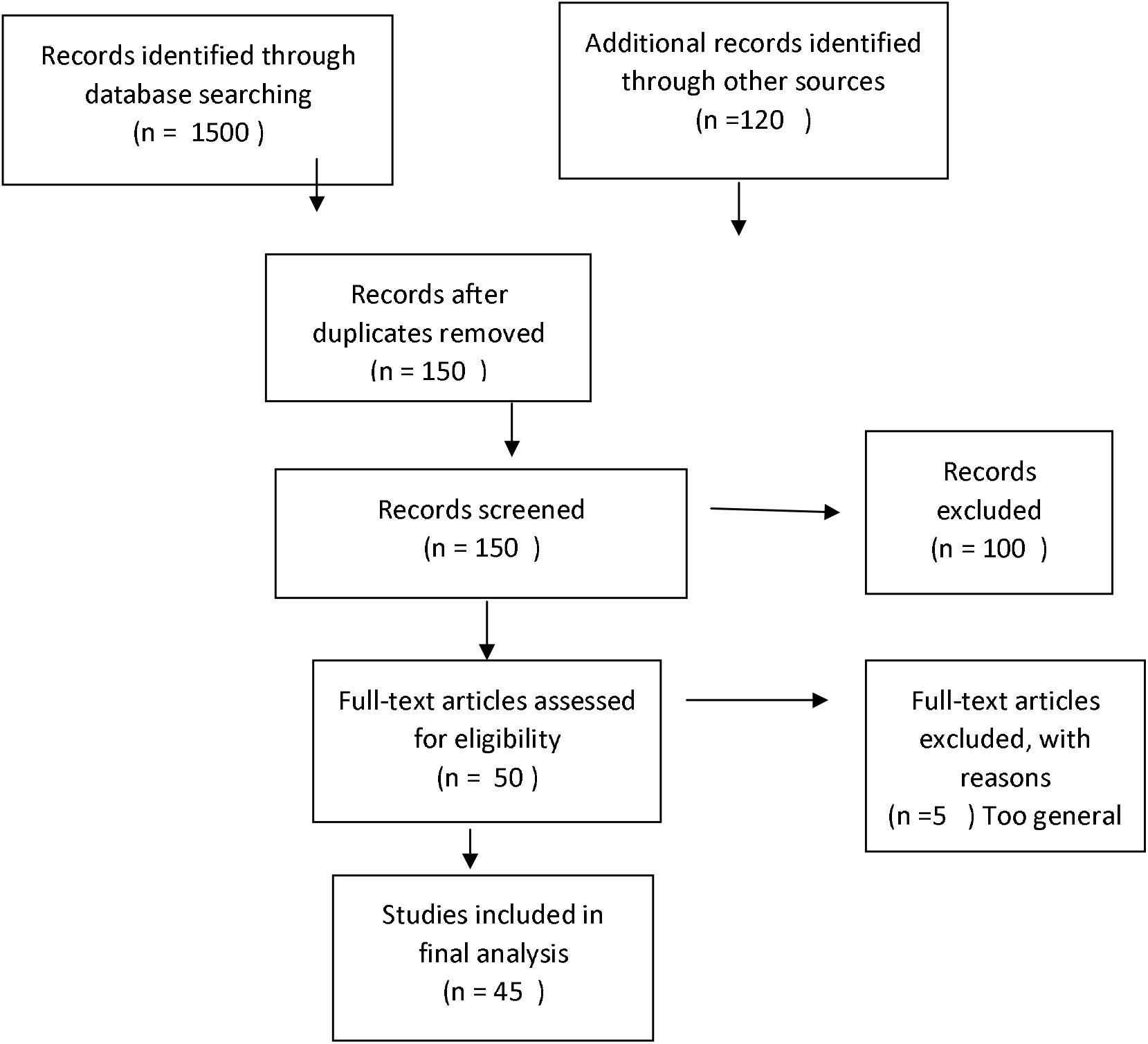
PRISMA flow chart for literature review.

The records identified through database searching were more than 1500; 120 were identified through other means, either books, thesis/dissertations or journals. After screening duplicates and irrelevant studies, 150 studies remained. From the remaining 150, a further 100 were either too general for area of focus or failed to access full texts (20). Finally, there were 50 full text articles, which were reviewed, and only 45 met the PICO criteria and were included for final synthesis. The following attributes were delineated from the literature: See table 1 below.

**Table 1:** Attributes of elements of women empowerment.

| **Element** | **Attributes** | **Key attributes** |
| --- | --- | --- |
| Role of women<br>(Person) | Adequate knowledge, encouraging, convincing, informed decisions, recipients of messages, facilitate couple communication, decision making, advising, educating, withholding sex, make VMMC precondition, influencing, refusal of VMMC, negotiating power, likely to persuade, talking to men, insisting on circumcision, persuasive power, negotiate condom use, influence men, make decisions, sway partners. | Knowledgeable, recipients of information, facilitate couple communication, encouraging, convincing, negotiating power, persuasive power, influencing |
| Promote VMMC uptake<br>(Nursing) | Women favour VMMC, involvement of women, source of information, women prefer circumcised men, influencing male interest, discussion with partner, readiness for MC, acceptance of MC, impact men's decision, ensure success of MC, accept MC from partner, influence men's decision making, overt or covert influence, convince men, high knowledge and acceptance of MC, availability, self-efficacy, social support, determinant of readiness, joint decision, duty to convince, narrows inequality, improves overall health, positive correlation | Involvement of women, women prefer circumcised men, source of information, influencing men, high knowledge and acceptance of VMMC |
| Barriers<br>(Negative) | Cultural beliefs, shame, older age, post-surgical abstinence, pain occurring during the | Fear of pain, myths and misconceptions, |
| environment) | procedure or after the procedure, myths and misconceptions, partner refusal, fear of HIV test, long healing period, VMMC not part of culture, fear of poor wound healing, fear of losing job, promiscuity, concern for safety, reduced penis sensitivity, adverse events, low risk perception. | post-surgical abstinence, adverse events, long healing period, fear of HIV test |
| Facilitators<br>(Health) | Reduction of HIV by 60%; reduction of STIs; eliminates occurrence of posthitis; phimosis and paraphimosis; reduces occurrence of UTI in infants; balanitis; penile cancer and cancer of the cervix; provided by well trained personnel in a safe environment; use of anaesthetic; hygiene; longer to ejaculate leading to sexual satisfaction; positive attitudes; religion; perceived attractiveness; belief the women prefer circumcised men; social support. | Reduction of HIV and STIs, reduction of cancer of the penis and cervix, sexual pleasure, hygiene. |

### Identification of concepts

The concepts were selected from literature search by searching for words and or group of words that represented the process of women empowerment as the key central concept and purpose of the study; according to Chinn & Kramer’s process of concept identification [10]. The concepts are change agent, recipient, procedures, dynamics and outcome. The main central concept are the women as the key social support system described in the formulated framework. See S1 Figure.

### Structural description of the framework

The structure of the women empowerment framework to promote VMMC uptake for HIV prevention is summarised in three main steps, all linked to the main central concept of women empowerment responsible for receiving VMMC knowledge, influencing positive perceptions to men and facilitating access of VMMC services to men. The peripheral concepts all linked to the central concept are health care workers as the change agents for accurate VMMC knowledge dissemination to women. This impact on effective VMMC information dissemination by women to men, causing positive attitudes towards VMMC and leading to enhanced VMMC practice and successful program implementation. See S2 figure.

## DISCUSSION

This study developed a women empowerment framework, from literature gathered on the role of women and women identified as significant elements within social support systems to promote VMMC uptake. Knowledgeable women have increased negotiating power and are capable to influence, teach, sway, persuade and convince men and boys about VMMC to promote VMMC uptake.

When women are educated, they have a higher power in negotiating skills and they become equal to men in decision-making [14]. They, therefore, are empowered to confidently negotiate for MC and condom use as preventive measures against sexually transmitted infections including HIV [14]. This is in support of the common saying, educate a woman, and you have educated a whole nation. Exploration of how increased knowledge can influence how women view MC is elaborated in the following studies. Findings of a study on effects of male circumcision on women were that having some formal education or training impacted on improved status of women. When a woman has a higher status, she can confidently manage any problems of MC and she is able to broadly choose options in her life. [15]. When one has power, they are able to decide and influence others on decisions. For women to have that power, it is crucial that they have a conducive environment and adequate knowledge to decide appropriately. The impact of power in women is shown in their ability to make choices without any fear of something bad to happen. It is important for women to have adequate knowledge for them to be able to contribute effectively in MC issues [16].

The empowerment involves social support empowerment in the five categories of information support, esteem support, emotional support, social network and tangible support [17]. Information support involves the women creating awareness and educating men about VMMC. The women give emotional support to their nervous and jittery partners and sons before and after the procedure through counselling and encouragement. Companionship support involves women influencing and persuading men to take up VMMC while social network support will be the support given to men by women through accompanying them for VMMC services and reviews. Tangible support will be women assisting men with wound care and abstinence post procedure.

Social support systems can make one more resilient in times of stress, provide encouragement and lower one’s stress level and feelings of loneliness. The role of women as key social support systems is important and positive for healthy individuals [13]. An effective social support system pre-MC reinforces, enhances or encourages men to go for MC [18]. Since the women are well empowered with adequate knowledge, they will be very effective in supporting men pre and post VMMC and giving them the correct care. The pre-procedure support by the women to men include information giving and encouraging men to go for the procedure. If women have adequate knowledge on MC, they are able to convince their husbands and sons [19]. Another study observed that in some cases, men had inadequate information on MC as compared to their wives causing women to be more influential to their partners on issues to do with MC. Some females in the same study believed they have a way of talking to their partners to accept male circumcision [20]. Acceptance of MC by men will cause increased VMMC uptake and prevalence, and achievement of VMMC targets. High VMMC prevalence was proven to reduce HIV prevalence in a bid towards creating an HIV free generation.

## SIGNIFICANCE AND RECOMMENDATIONS

The framework provides guidance in VMMC services from educational interactions among health care workers, women and men to demand creation approaches utilizing the women as partners and mothers of eligible men for VMMC.

The framework has not yet been tested on a broad scale; hence, an area of recommendation for future research to further validate the framework on a broader scale; the researcher intends to conduct it as a postdoctoral project.

It is hypothesized that a highly empowered woman becomes positive about VMMC hence can influence men to take up the program.

The framework needs to be adopted and adapted for other men related public health programs e.g. cancer of prostrate awareness, HTS campaigns etc. and assess its applicability.

Health system administrators especially in male circumcision are recommended to seriously consider use of this formulated framework to incorporate women as key change agents in VMMC uptake. This can be initiated by demand creation policy revision to acknowledge the importance of women in VMMC uptake promotion.

## Data Availability

All required data will be provided by the corresponding author

## SUMMARY

Several studies support the important role of women as significant social support systems in influencing men to undergo male circumcision [3, 4, 7, 8, 13 & 15]. These studies and various other studies and literature led the researchers to conduct the study to develop a women empowerment framework focusing on women as the recipients of VMMC information and influencers in promoting VMMC uptake. The researchers used a theory-generative design to explore literature, develop and describe a women empowerment framework for promoting VMMC uptake. A model to promote VMMC uptake formed the basis of this framework [18]. The three-step framework is expected to direct demand creation approaches by women; and improve VMMC uptake in Zimbabwe.

## Acknowledgements

The authors express sincere gratitude to the University of South Africa for the academic support, and the MRCZ for the ethical clearance; children, Chido, Tendai and Vimbai for their unconditional support and encouragement and colleague, ‘Mambo’ Morgen Chinoda for all the IT assistance and VMMC updates. Most of all, we give glory to God, for the gift of health and life to be able to go through the project.

## Author contributions

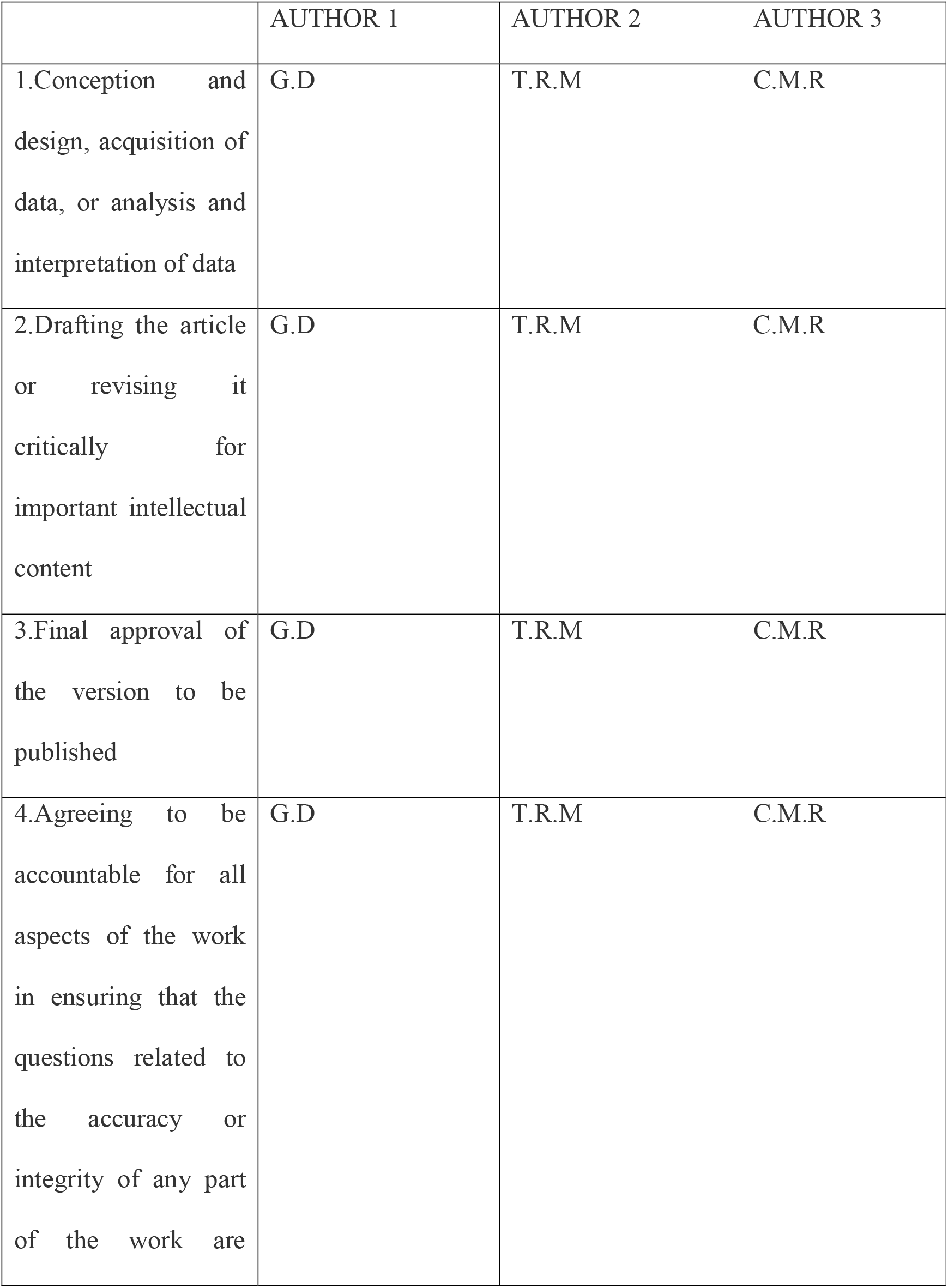
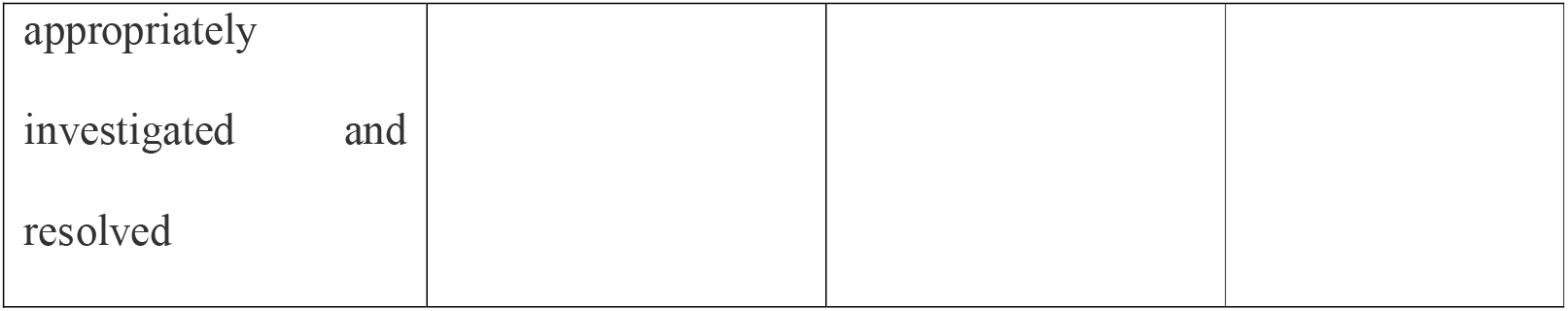

## Supporting information

**Figure 1:**
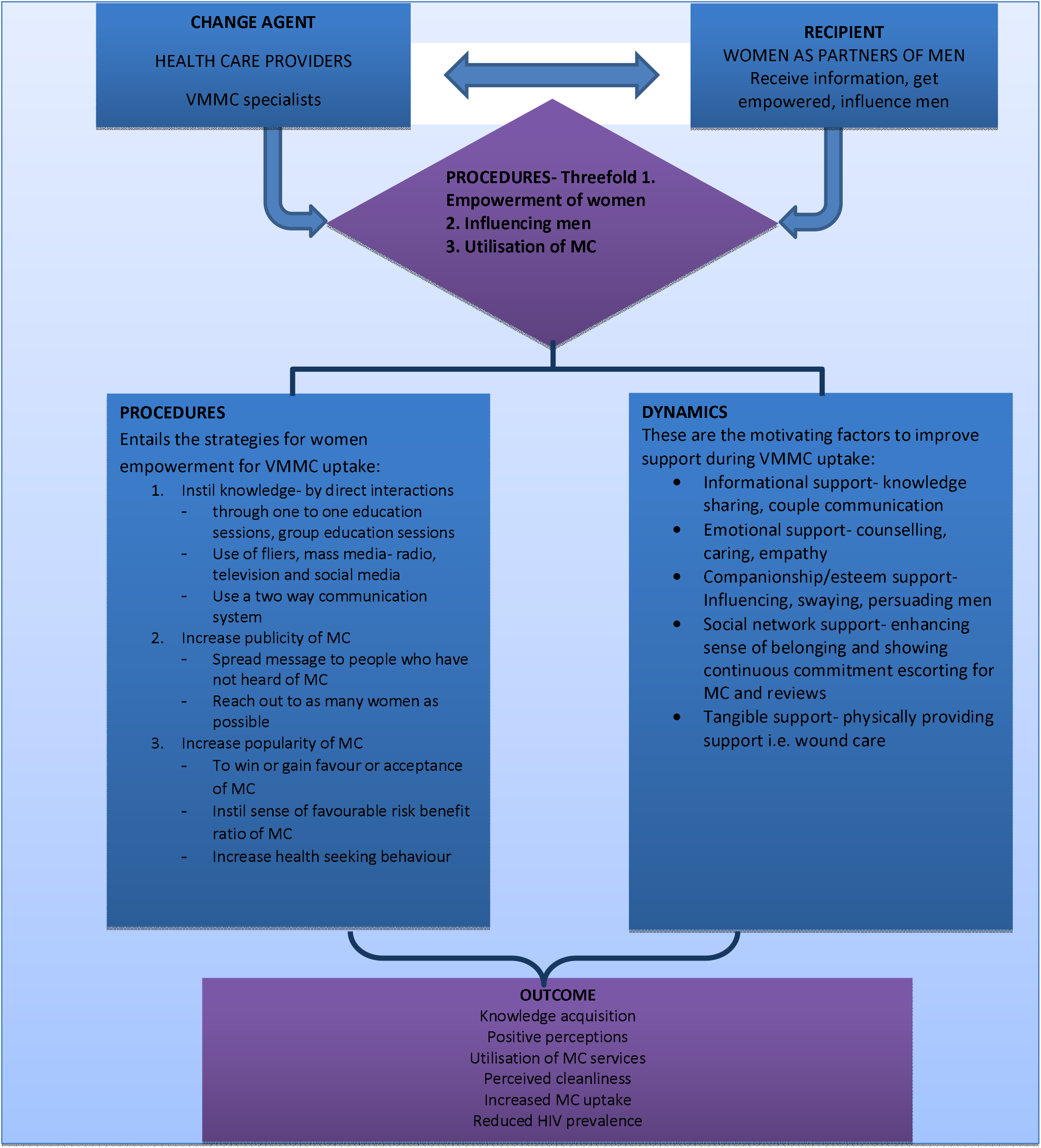
Relationship of Concepts diagram.

**Fig 2:**
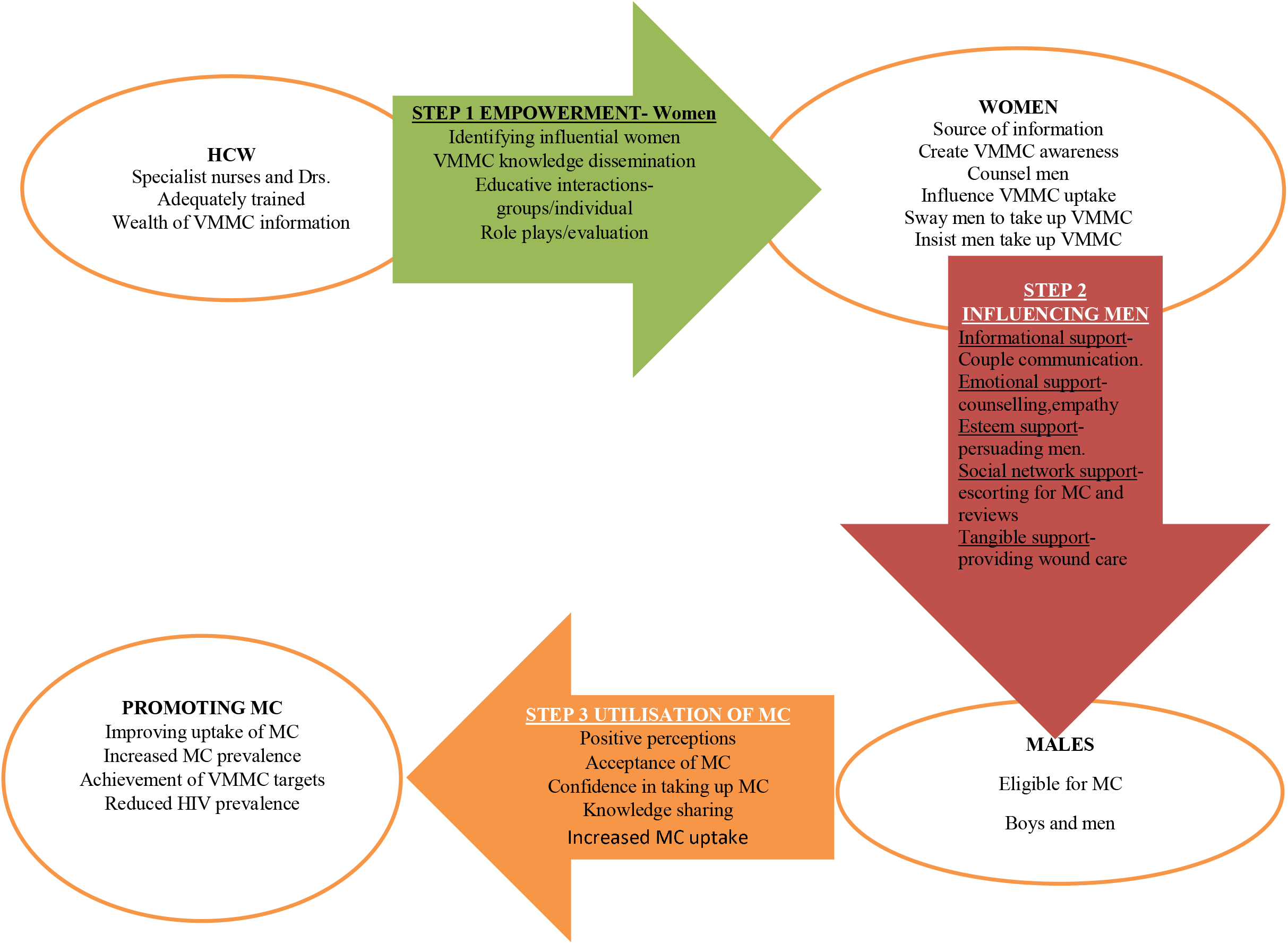
Women empowerment framework.

